# *FAT4* Identified as a Potential Modifier of Orofacial Cleft Laterality

**DOI:** 10.1101/2021.03.16.21253740

**Authors:** Sarah W. Curtis, Daniel Chang, Miranda R. Sun, Michael P. Epstein, Jeffrey C. Murray, Eleanor Feingold, Terri H. Beaty, Seth M. Weinberg, Mary L. Marazita, Robert J. Lipinski, Jenna C. Carlson, Elizabeth J. Leslie

## Abstract

Orofacial clefts (OFCs) are common (1 in 700 births) congenital malformations that include cleft lip (CL) and cleft lip and palate (CLP). These OFC subtypes are also heterogeneous themselves, with the cleft lip occurring on the left, right, or both sides of the upper lip. Unilateral CL and CLP have a 2:1 bias towards left-sided clefts, suggesting a nonrandom process. Here, we performed a study of left- and right-sided clefts within the CL and CLP subtypes to better understand the genetic factors controlling cleft laterality. We conducted genome-wide modifier analyses by comparing cases that had right unilateral CL (RCL; N=130), left unilateral CL (LCL; N=216), right unilateral CLP (RCLP; N=416), or left unilateral CLP (LCLP; N=638), and identified a candidate region on 4q28, 400 kb downstream from *FAT4*, that approached genome-wide significance for LCL vs. RCL (p = 8.4×10^−8^). Consistent with its potential involvement as a genetic modifier of cleft lip, we found that *Fat4* exhibits a specific domain of expression in the mesenchyme of the medial nasal processes that form the median upper lip. Overall, these results suggest that the epidemiological similarities in left-to right-sided clefts in CL and CLP are not reflected in the genetic association results.

## Introduction

Orofacial clefts (OFCs) - cleft lip (CL), cleft lip with palate (CLP), and cleft palate (CP) - are common congenital craniofacial anomalies that affect 1 in 700 births globally (Rahimov, Jugessur, & Murray, 2012). OFCs lead to feeding difficulties and require multiple surgeries early in life to repair (Wehby & Cassell, 2010; Wehby et al., 2006). In addition to these early childhood complications, affected individuals are at a higher risk for dental (Akcam, Evirgen, Uslu, & Memikoğlu, 2010; Lourenco Ribeiro, Teixeira Das Neves, Costa, & Ribeiro Gomide, 2003; Ribeiro, das Neves, Costa, & Gomide, 2002) and speech problems (Jocelyn, Penko, & Rode, 1996; Mort, 1968), ear infections (Jocelyn et al., 1996; Nackashi, Dedlow, & Dixon-Wood, 2002), various forms of cancer (Dietz et al., 2012; Menezes et al., 2009), and mental health concerns (Christensen & Mortensen, 2002), and these individuals have an overall higher rate of mortality across the life span (Christensen, Juel, Herskind, & Murray, 2004). Beyond its impact on health, individuals with OFCs also incur additional financial burdens, with the lifetime costs for cleft-related surgeries, hospital stays, orthodontic treatments, and speech therapy exceeding $200,000 (Wehby & Cassell, 2010; Wehby et al., 2006).

Although the exact causal mechanism of OFC formation is unknown, OFC pathogenesis is multifactorial, and there is evidence for both genetic and environmental factors contributing to OFC risk (Leslie & Marazita, 2013). This was supported both by population-based studies showing increased risk of OFC in individuals with a family history of OFCs compared to the general population (Sivertsen et al., 2008) and twin studies showing a higher OFC concordance among monozygotic twins than dizygotic twins (Little & Bryan, 1986). More recently, genome-wide association studies (GWAS) have identified 40-50 different genomic regions associated with OFCs, such as the association between OFCs and *IRF6* (Terri H Beaty et al., 2010; Stefanie Birnbaum et al., 2009), *MAFB* (Terri H Beaty et al., 2010; Lennon et al., 2012), *ARHGAP29* (Terri H Beaty et al., 2010; Kerstin U Ludwig et al., 2012), 8q24 (Stefanie Birnbaum et al., 2009), and 1q32 (Terri H Beaty et al., 2010).

Genetic studies often group the CL and CLP OFC subtypes into cleft lip with or without cleft palate (CL/P) based on several shared observations: (1) CL and CLP share a defect of the lip which forms prior to palatal development, (2) there is a 2:1 ratio of left- and right-sided clefts in both CL and CLP (K. K. H. Gundlach & C. Maus, 2006), and (3) the recurrence risk for CL and CLP are similar and both can occur within the same family. Although combining multiple subtypes can increase statistical power, doing so also dilutes any signal coming from one subtype. Recent genetic studies have demonstrated classifying OFCs by phenotypic characteristics, such as where the cleft occurs and the severity or laterality of the cleft lip, to create more homogenous subgroups can facilitate novel gene discovery (Carlson et al., 2019; Carlson, Standley, et al., 2017; Carlson, Taub, et al., 2017; Huang et al., 2019). One approach is to conduct a case-only modifier analysis, comparing two distinct cleft subtypes to each other, in order to identify genetic modifiers for OFC phenotypes. We previously used this approach to identify a locus on 16p21 that increases risk of CL versus CLP (Carlson, Standley, et al., 2017), a locus near *PAX1* that increases risk of bilateral CL versus unilateral CL (Curtis et al.), and clusters of rare variants near *IRF6* associated with unilateral versus bilateral CL/P (Carlson, Taub, et al., 2017). The findings from the previous studies combined with the non-random distribution suggests that there may be underlying genetic factors that contribute to phenotypic differences among OFC cases and grouping these subtypes together could potentially overlook genetic factors unique to the pathogenesis of a specific subtype. Importantly, the genetic risk of individual subtypes of OFCs, the preponderance of left-versus right-sided clefts is still not well understood.

To better understand the genetic variants associated with the side of unilateral CL/Ps, we conducted both subtype-specific GWAS and genome-wide modifier scans comparing left- and right-sided unilateral clefts (CL/P) and within the CL and CLP groups separately. Finally, we sought to better understand the genetic differences between left and right unilateral clefts by using model systems and publicly available databases to functionally annotate any associated loci.

## Methods

### Study population

Samples were obtained from a multi-ethnic cohort from the Pittsburgh Orofacial Cleft (POFC) study, with recruitment occurring in 18 OFC treatment centers in North, Central, and South America, Asia, and Europe, as part of genetic studies by the University of Pittsburgh Center for Craniofacial and Dental Genetics and the University of Iowa (Leslie, Carlson, et al., 2016; Leslie, Liu, et al., 2016). Recruitment was approved by the IRB of each recruiting site, as well as the IRB of the University of Pittsburgh and University of Iowa, and written, informed consent was obtained for each research subject (Leslie, Carlson, et al., 2016; Leslie, Liu, et al., 2016). Eligibility was determined by whether or not the individual had OFC and the availability of details of the OFC type. Cases for analyses were selected from unrelated individuals who had cleft lip only (CL) or cleft lip and palate (CLP). For this study, cases were classified as having either a left unilateral CL (LCL, N = 216), a right unilateral CL (RCL, N = 130), a left unilateral CLP (LCLP, N = 638), or a right unilateral CLP (RCLP, N = 416). Controls were defined as individuals unrelated to cases who have no known history of OFC or other craniofacial anomalies (N = 1626).

### Genotyping quality control

Samples were genotyped for a combination of 580,000 single nucleotide polymorphic (SNP) markers using the Illumina HumanCore+Exome platform, and 15,980 SNPs in candidate genes and loci identified by previous studies of OFCs (Leslie, Liu, et al., 2016). The dataset analyzed in this study underwent quality control (QC) using pipelines developed by the University of Washington Genetics Coordinating Center as described previously (Laurie et al., 2010; Leslie, Carlson, et al., 2016; Leslie, Liu, et al., 2016). This process involved examining samples for duplicates, batch effects, chromosomal anomalies, familial relatedness, Mendelian errors among relatives, and population structure. SNP probe quality was also inspected by examining inter-sample comparisons, missing call rates, separation of clusters during genotype calling, and deviations from Hardy-Weinberg equilibrium. After QC, the final number of genotyped SNPs was 539,473, with 293,633 SNPs having a minor allele frequency 1% or greater (Leslie, Carlson, et al., 2016; Leslie, Liu, et al., 2016). This data was then used to impute additional SNPs using the IMPUTE2 software with the 1000 Genome Projects (phase 3) as the reference panel, with haplotypes created using the SHAPEIT2 software (Leslie, Carlson, et al., 2016; Leslie, Liu, et al., 2016). The most-likely genotypes were selected for statistical analysis if the highest probability (r^2^) > 0.9. SNPs showing deviations from Hardy-Weinberg equilibrium in European controls (p < 1 × 10^−4^), a MAF < 5%, or imputation INFO scores < 0.5 were excluded from downstream analyses.

### Subtype-specific GWAS

A single-variant GWAS was done, comparing each subtype to the unrelated controls. Association between subtypes and genetic variants that passed QC was tested using a logistic regression model controlling for sex and 18 principal components (PCs) to adjust for genetic ancestry as implemented in PLINK v1.9 (Chang et al., 2015; Purcell S, 2007). P-values less than 5 × 10^−8^ were considered genome-wide significant, and associations were considered suggestive if the p-value was between 1 × 10^−5^ and 5 × 10^−8^. Regional association plots were created using LocusZoom (Randall J Pruim et al., 2010)

### Modifier GWAS

In addition to the subtype-specific analysis, we conducted a case vs. case modifier GWAS to identify SNPs associated with differences between OFC subtypes. In contrast to traditional analyses comparing separate analyses of cases vs. controls, the modifier approach has high statistical power to find genetic risk factors that differ between the two groups, but no power to find factors equally associated with both groups (Yang, Lee, Goddard, & Visscher, 2011). In this approach, LCL were compared RCL, LCLP were compared to RCLP, and LCL/P were compared to RCL/P. Similar to the analysis above, this was done using a logistic regression model controlling for sex and 18 principal components (PCs) to adjust for genetic ancestry as implemented in PLINK v1.9 (Chang et al., 2015; Purcell S, 2007).

### Comparisons between CL and CLP subtype-specific and modifier analyses

For the subtype-specific and the modifier analyses, the odds ratios and 95% confidence intervals for SNPs that were suggestive (p < 1 × 10^−5^ in either analysis) in a pair of analyses were compared to see if they either overlapped or were in similar directions. Pairs of analyses included different cleft sides within a cleft subtype (e.g., LCL vs. RCL) and the same side between subtypes (e.g., LCL vs. LCLP). In order to determine whether the SNPs associated with individual subtypes were novel compared to what has already been associated with CL/P, SNPs that were within 50kb of SNPs previously associated with OFCs (Carlson et al., 2019; Huang et al., 2019; Leslie, Carlson, et al., 2016; Yu et al., 2017) were identified.

### Fat4 gene expression analysis

To determine biological relevance of the 4q28 locus, the expression of the *Fat4* candidate gene was characterized by *in situ* hybridization (ISH) of mouse embryos. Studies involving mice were conducted in strict accordance with the recommendations in the National Institutes of Health’s “Guide for the Care and Use of Laboratory Animals.” The protocol was approved by the University of Wisconsin-Madison School of Veterinary Medicine Institutional Animal Care and Use Committee (protocol number 13–081.0). C57BL/6J mice (*Mus musculus*) were purchased from The Jackson Laboratory. Timed pregnancies were established as described previously (Heyne et al., 2015). Embryos at indicated gestational days were dissected in PBS and fixed in 4% paraformaldehyde in PBS overnight followed by graded dehydration (1:3, 1:1, 3:1 volume per volume [v/v]) into 100% methanol for storage. For sections, embryos were rehydrated and embedded in 4% agarose gel, and 50 μm coronal sections were made with a vibrating microtome. ISH was performed on whole-mount embryos and sections as previously described (Heyne et al., 2016). *Fat4* ISH riboprobe primers were designed with IDT PrimerQuest and affixed with the T7 polymerase consensus sequence plus a 5-bp leader sequence to the reverse primer (forward primer: CAGAGAGCAGAGGTAGAAATAAC, reverse primer with T7 consensus and leader sequence underlined: <u>CGATGTTAATACGACTCACTATAGGG</u>CTGAGAGTTGACACCATCATC). A MicroPublisher 5.0 camera connected to an Olympus SZX-10 stereomicroscope was used for imaging.

### Replication cohort

To replicate any statistically significant results from the modifier analysis, we analyzed genetic data from the GENEVA International Cleft Consortium (T. H. Beaty et al., 2010; Carlson et al., 2019; Leslie, Carlson, et al., 2016). Briefly, this cohort recruited case-parent trios, where the affected individual had cleft lip with or without cleft palate. The samples were genotyped for approximately 589K SNPs using the Illumina Human610-Quadv.1_B BeadChip. This data was then phased using SHAPEIT, and imputation was performed using IMPUTE2 to the 1000 Genomes Phase I (June 2011) reference panel. Imputed genotype probabilities were converted to most-likely genotype calls with GTOOL (http://www.well.ox.ac.uk/~cfreeman/software/gwas/gtool.html). This dataset was subsequently filtered to only include SNPs with a MAF > 5%. Individuals that were genotyped as part of both the POFC study and the GENEVA study were removed from the replication cohort so that the two groups would be independent. Only the cases from the non-overlapping GENEVA cohort were selected, and were classified as having either a LCL (N = 219), RCL (N = 107), LCLP (N = 428), or RCLP (N = 250). Principal components (PCs) of ancestry were calculated using PLINK (v1.9) (Chang et al., 2015). Modifier analyses (comparing LCL to RCL and LCLP to RCLP) were conducted using logistic regression models in PLINK (v1.9), with sex and the first 4 PCs as covariates. Because of the differences in genotyping arrays and imputation panels, we used a region-based replication strategy, where we tested the association of SNPs in the same region as the associated SNP. To account for multiple testing, we used a Bonferroni correction for the number of SNPs in the region (0.05/the number of SNPs tested).

### Epigenomic context of results

Functional enrichment of epigenetic marks was tested by first annotating the SNPs to the craniofacial functional regions defined by Wilderman *et. al*. (Wilderman, VanOudenhove, Kron, Noonan, & Cotney, 2018) for human embryos at CS13, CS14, CS15, CS17, and CS20 (4.5-8 weeks post conception). Enrichment tests were done using a chi-square test with the top SNPs (p < 1 × 10^−3^) for both modifier analyses and each subtype analysis, and calculated odds ratios and their 95% CI.

### Gene set enrichment test

We tested for enrichment of SNPs from each modifier and subtype-specific analysis in genes associated with other laterality disorders using MAGMA (v1.08)(de Leeuw, Mooij, Heskes, & Posthuma, 2015). The list of phenotypes and genes included genes involved with visceral asymmetry from the mouse phenotype database and primary ciliary disease in humans, as described in de Kovel *et al*. (de Kovel & Francks, 2019).

### Genetic interaction with *FAT4*

We tested for genetic interaction in the modifier analyses between SNPs in the *FAT4* gene and known OFC regions (any SNP within 50kb of a known OFC locus) (Carlson et al., 2019; Huang et al., 2019; Leslie, Carlson, et al., 2016; Yu et al., 2017) using PLINK v1.9 (Chang et al., 2015; Purcell S, 2007), controlling for sex and 18 principal components (PCs) to adjust for genetic ancestry. Interaction was tested in LCL, RCL, LCLP and RCLP compared to controls separately. A Bonferroni correction for the number of regions (N = 1,291) was used for statistical significance.

## Results

### Subtype-specific GWAS

We performed a genome-wide subtype-specific analysis to identify genetic variants that may distinguish between left- and right-sided unilateral clefts in cleft lip only (CL) and cleft lip and palate (CLP) subtypes. Our analyses included 216 participants with LCL, 130 with RCL, 638 with LCLP, and 416 with RCLP. While these subjects were not a population-based sample, the relative frequencies of left- and right-sided OFCs in our sample were consistent with published population based frequencies (Table S1) (K. K. Gundlach & C. Maus, 2006). Cases of each subtype group were separately compared to 1,626 unaffected controls. This analysis has the potential to detect variants that increase risk for an OFC in general or variants that increase risk in just one or more subtypes. As expected, the region on 8q24, a well-established association with risk to both CL and CLP, was the only region to reach genome-wide significance in any analysis and many of the suggestive loci overlapped previously reported CL/P loci (Figure S1-S2; Table S2-S5) (T. H. Beaty et al., 2010; S. Birnbaum et al., 2009; Leslie, Carlson, et al., 2016; Mangold et al., 2010; Nikopensius et al., 2009). However, several loci not been previously implicated in OFCs achieved suggestive significance, such as 16q23.2 for LCL (lead SNP: rs16953809; p = 1.43 × 10^−7^), 7p12 for RCL (lead SNP: rs138411667; p = 4.95 × 10^−7^), 4q35 for LCLP (lead SNP: rs4069861; p = 6.19 × 10^−7^), and 10p12 for RCLP (lead SNP: rs2497818; p = 1.57 × 10^−6^; Figure S3).

To determine whether these suggestive loci from the case-control analyses were similar across subtypes, we compared the odds ratios for SNPs that were suggestive in any of the four analyses. We compared these SNPs in four sets of different pairs to distinguish differences between the side of a cleft (i.e., LCL vs. RCL SNPs) and differences between cleft types (i.e. LCL vs. LCLP SNPs) (Figure 1). The SNPs apparently influencing risk to LCLP and RCLP were the most similar to each other, with 90.15% (N = 384) of SNPs having overlapping confidence intervals. The SNPs influencing risk for LCL and RCL were the most different with only 62.3% (N = 137) of SNPs having overlapping confidence intervals. When compared across cleft type, 76.9% of suggestive SNPs for LCL and LCLP and 72.4% for RCL and RCLP had overlapping confidence intervals. Across all comparisons, the SNPs with different ORs between subtypes were less likely to be the SNPs associated with OFCs in previous GWAS (p < 0.001; Table S6). These findings suggest CL and CLP have some shared risk alleles but also distinct risk alleles, which may independently affect the side of the cleft.

**Figure 1:**
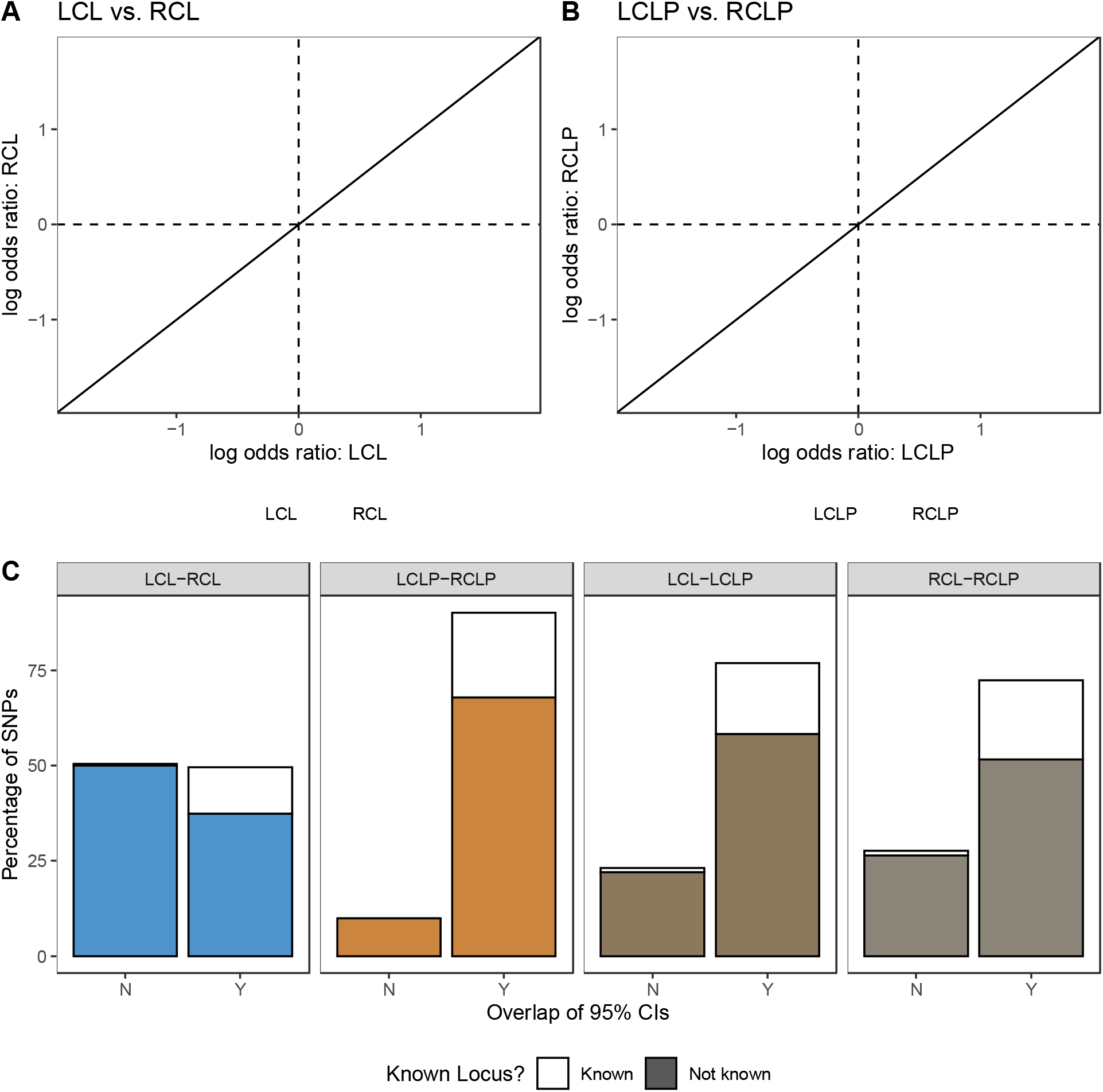
The odds ratio for SNPs that were suggestive (p < 1 × 10^−5^) or significant (p < 5 × 10^−8^) in the subtype-specific case-control analyses in were compared between right and left unilateral CL (A), right and left unilateral CLP (B), and were classified by whether the confidence interval for the odds ratio overlapped and whether the variant was known (C).

### Modifier GWAS

To further investigate the genetic differences within each subtype and disentangle the effects on side of the cleft from subtype, we next conducted a genome-wide modifier analysis. Because this is a case-to-case comparison, this analysis would be able to detect variants important for the formation of one type of cleft compared to other, but cannot detect variants that are generally influencing risk to both OFC subtypes in any comparison. We first compared LCL to RCL and LCLP to RCLP. In the LCL-RCL analysis, no region achieved genome-wide significant, but a locus on chromosome 4q28 approached this threshold (lead SNP: rs6855309; OR = 3.50; 95% CI: 2.21-5.54; p-value = 8.4 × 10^−8^; Figure 2A; Figure S4A; Table 1), and 11 other loci reached suggestive significance. In the LCLP-RCLP analysis, again no region reached genome-wide significance and 11 loci yielded suggestive significance (Figure 2B; Figure S4B; Table 2). Interestingly, when CL and CLP were combined into CL/P (as typical in genetic studies of OFCs) the LCL/P vs RCL/P modifier analysis again failed to identify any genome-wide significant SNPs, and only 12 loci reached suggestive significance (Figure S5; Table S7), indicating a lack of power or the possibility of heterogeneity in genetic factors predisposing to the side of a cleft in CL versus CLP.

**Figure 2:**
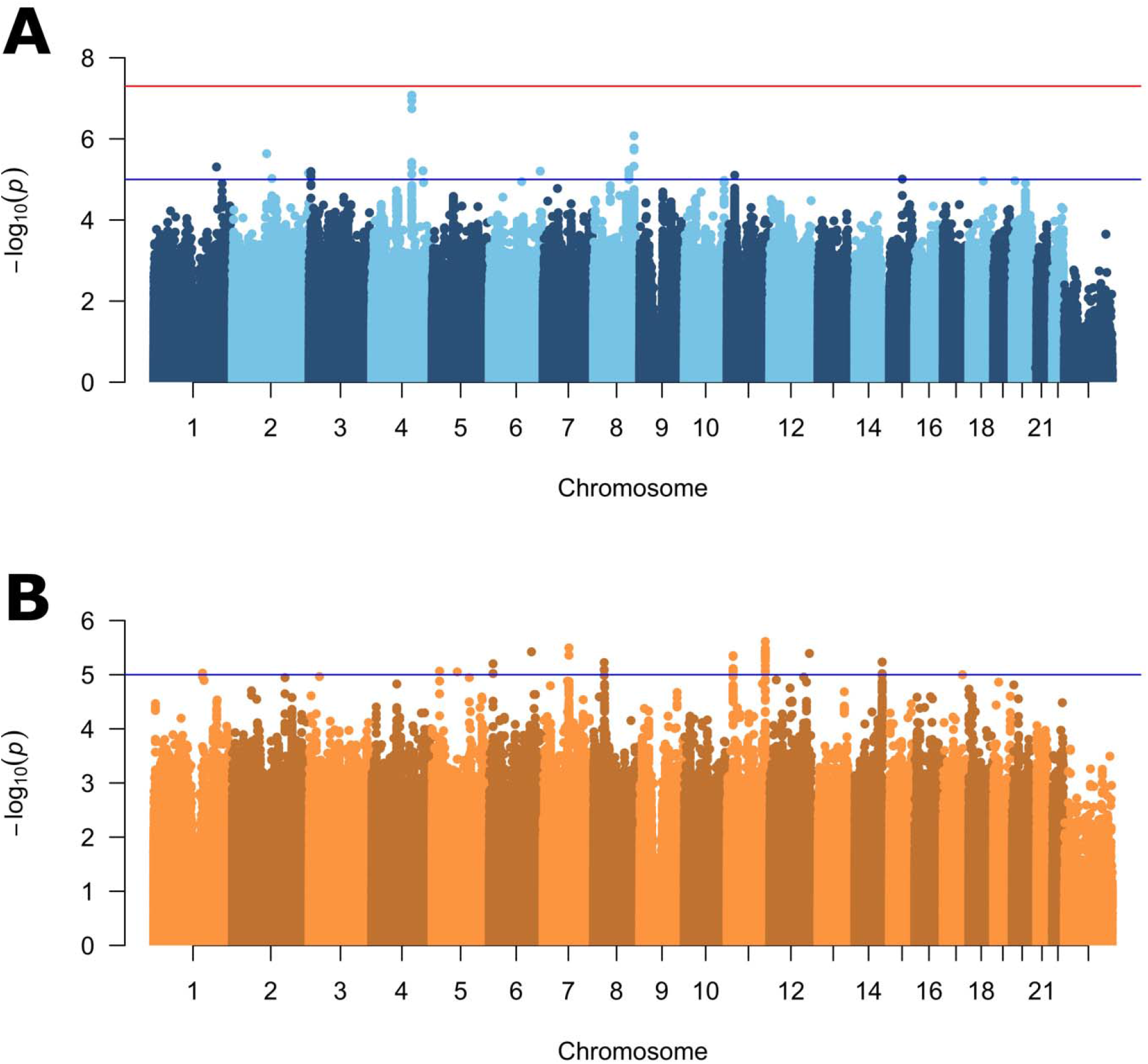
Manhattan plots of –log_10_(p-values) from the left unilateral vs right unilateral modifier analysis in participants with cleft lip (A), and cleft lip and palate (B). Lines indicate suggestive (blue) and genome-wide (red) thresholds for statistical significance. The genomic inflation factors are 1.08 and 1.02, respectively.

**Table 1:** Suggestive modifiers for right unilateral and left unilateral CL

| **Locus** | **Lead SNP** | **Reference allele** | **Alternate allele** | **AF<sup>1</sup>** | **Odds Ratio** | **95% CI** | **P-value** |
| --- | --- | --- | --- | --- | --- | --- | --- |
| 4q28.1 | rs6855309 | C | T | 0.29 | 3.50 | (2.21, 5.54) | $8.40 \times 10^{-8}$ |
| 8q24.21 | rs13267780 | G | A | 0.21 | 0.32 | (0.21, 0.51) | $8.40 \times 10^{-7}$ |
| 2q12.3 | rs139260643 | C | T | 0.28 | 3.30 | (2.01, 5.43) | $2.32 \times 10^{-6}$ |
| 1q32.1 | rs12732777 | C | A | 0.63 | 0.36 | (0.23, 0.56) | $4.93 \times 10^{-6}$ |
| 8q23.3 | rs62520628 | C | T | 0.32 | 2.91 | (1.83, 4.64) | $5.91 \times 10^{-6}$ |
| 4q32.2 | rs5010472 | A | G | 0.61 | 2.31 | (1.61, 3.33) | $6.12 \times 10^{-6}$ |
| 6q26 | rs71004034 | A | AT | 0.26 | 0.37 | (0.24, 0.57) | $6.25 \times 10^{-6}$ |
| 3p26.1 | rs58292735 | G | A | 0.28 | 0.40 | (0.27, 0.60) | $6.38 \times 10^{-6}$ |
| 2q37.3 | rs12465491 | C | T | 0.29 | 0.39 | (0.26, 0.59) | $6.96 \times 10^{-6}$ |
| 11p14.3 | rs10606454 | ATTTT | A | 0.45 | 0.39 | (0.26, 0.59) | $7.89 \times 10^{-6}$ |
| 2q14.3 | rs2553629 | A | T | 0.53 | 2.42 | (1.64, 3.59) | $9.52 \times 10^{-6}$ |
| 15q22.2 | rs112409955 | CAAAT | CAAATAAAT | 0.26 | 0.38 | (0.24, 0.58) | $9.82 \times 10^{-6}$ |

<sup>1</sup>Allele frequency of alternate allele

**Table 2:** Suggestive modifiers for right unilateral and left unilateral CLP

| **Locus** | **Lead SNP** | **Reference allele** | **Alternate allele** | **AF<sup>1</sup>** | **Odds Ratio** | **95% CI** | **P-value** |
| --- | --- | --- | --- | --- | --- | --- | --- |
| 11q23.3 | rs499804 | C | A | 0.22 | 0.59 | (0.47, 0.73) | $2.45 \times 10^{-6}$ |
| 7q21.11 | rs2194751 | G | A | 0.48 | 1.55 | (1.29, 1.87) | $3.20 \times 10^{-6}$ |
| 6q23.2 | rs17642884 | A | G | 0.08 | 0.46 | (0.33, 0.64) | $3.79 \times 10^{-6}$ |
| 12q24.31 | rs34152756 | C | CA | 0.49 | 1.69 | (1.35, 2.11) | $4.05 \times 10^{-6}$ |
| 11q15.1 | rs112004298 | C | T | 0.09 | 2.32 | (1.62, 3.33) | $4.45 \times 10^{-6}$ |
| 14q32.33 | rs34414198 | C | A | 0.57 | 0.10 | (0.04, 0.28) | $5.85 \times 10^{-6}$ |
| 8p12 | rs10588090 | TAGTA | T | 0.35 | 0.63 | (0.51, 0.77) | $5.98 \times 10^{-6}$ |
| 6p24.1 | rs9471440 | T | C | 0.17 | 0.56 | (0.43, 0.72) | $6.29 \times 10^{-6}$ |
| 5p14.2 | rs1568458 | T | A | 0.71 | 1.62 | (1.31, 2.01) | $8.62 \times 10^{-6}$ |
| 5q14.1 | rs34586243 | GA | G | 0.70 | 0.51 | (0.38, 0.69) | $8.91 \times 10^{-6}$ |
| 1q21.3 | rs10908436 | A | G | 0.57 | 1.71 | (1.35, 2.17) | $9.38 \times 10^{-6}$ |

<sup>1</sup>Allele frequency of alternate allele

Even though there were no genome-wide significant loci found in these modifier analyses, the lack of genome-wide significant loci in the LCL/P vs RCL/P analysis suggested that the modifiers for CL may differ (or have different directions of effect) than those for CLP. To test this hypothesis, we compared suggestive SNPs (p < 1 × 10^−5^) in either the LCL-RCL or LCLP-RCLP modifier analyses. Notably, there was no overlap between the SNPs that were suggestive in CL and the SNPs that were suggestive in CLP. Moreover, the odds ratio for any of suggestive SNPs in CL had an odds ratio near one in CLP and *vice versa* (Figure 3). This stark contrast in effect sizes suggests that the potential genetic modifiers of laterality in CL are distinct from those that are potential laterality modifiers in CLP.

**Figure 3:**
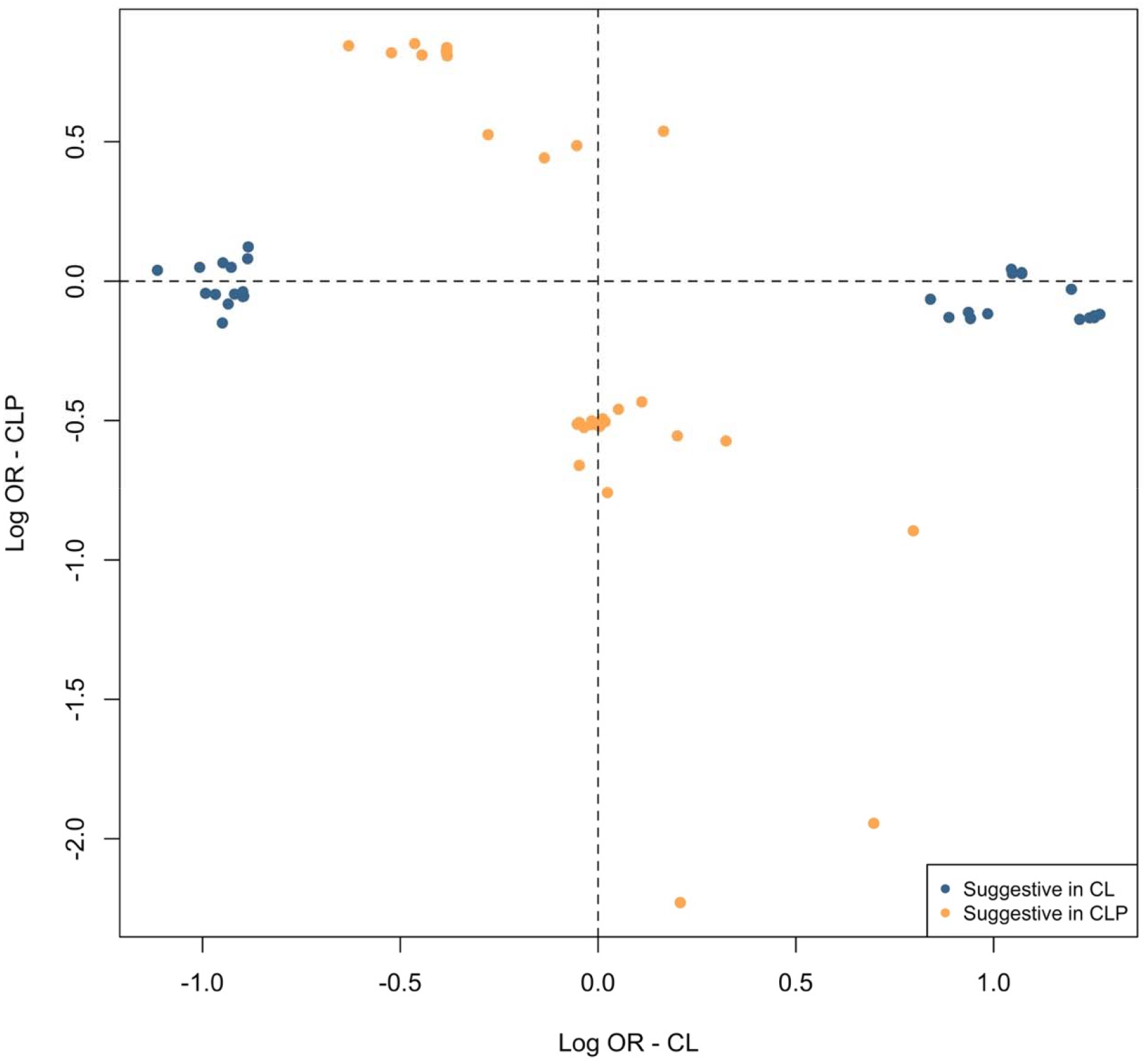
The odds ratio for SNPs yielding suggestive significance (p < 1 × 10^−5^) in the modifier analysis in CL or CLP were compared. No SNPs achieved suggestive significance in both CL and CLP.

### Attempted replication and Fat4 gene expression analysis

Given the LCL-RCL modifier top hits did not overlap with the LCLP-RCLP top hits, we next focused on the best modifier candidate from these analyses. The associated SNPs in the top candidate locus from the LCL-RCL modifier analysis is located 400 kb downstream of *FAT4*, a gene involved in the regulation of planar cell polarity (Figure 4A). For the lead SNP (rs6855309), the minor allele increased risk for LCL over RCL (OR = 2.21) and no effect was seen in the LCLP-RCLP comparison (OR = 0.88), and increased risk for LCL (OR = 1.84) but decreased risk for RCL (OR = 0.64) in analyses versus controls (Figure 4B). We attempted to replicate this locus in an independent cohort, but none of the SNPs in the region were significant after considering multiple testing corrections in either CL (Table S8) or CLP (Table S9). To test whether it was biologically plausible this gene could be affecting OFC subtypes, expression of *Fat4* was examined during orofacial development in mice. We found that *Fat4* is expressed in the mesenchyme of the medial nasal processes that form the median aspect of the upper lip and primary palate. Expression was detected from gestation day 10.75 to 11.75, a period in which the medial nasal processes fuse with the adjacent processes to close the upper lip (Figure 5), indicating that *Fat4* is expressed at a time and location relevant for cleft lip formation.

**Figure 4:**
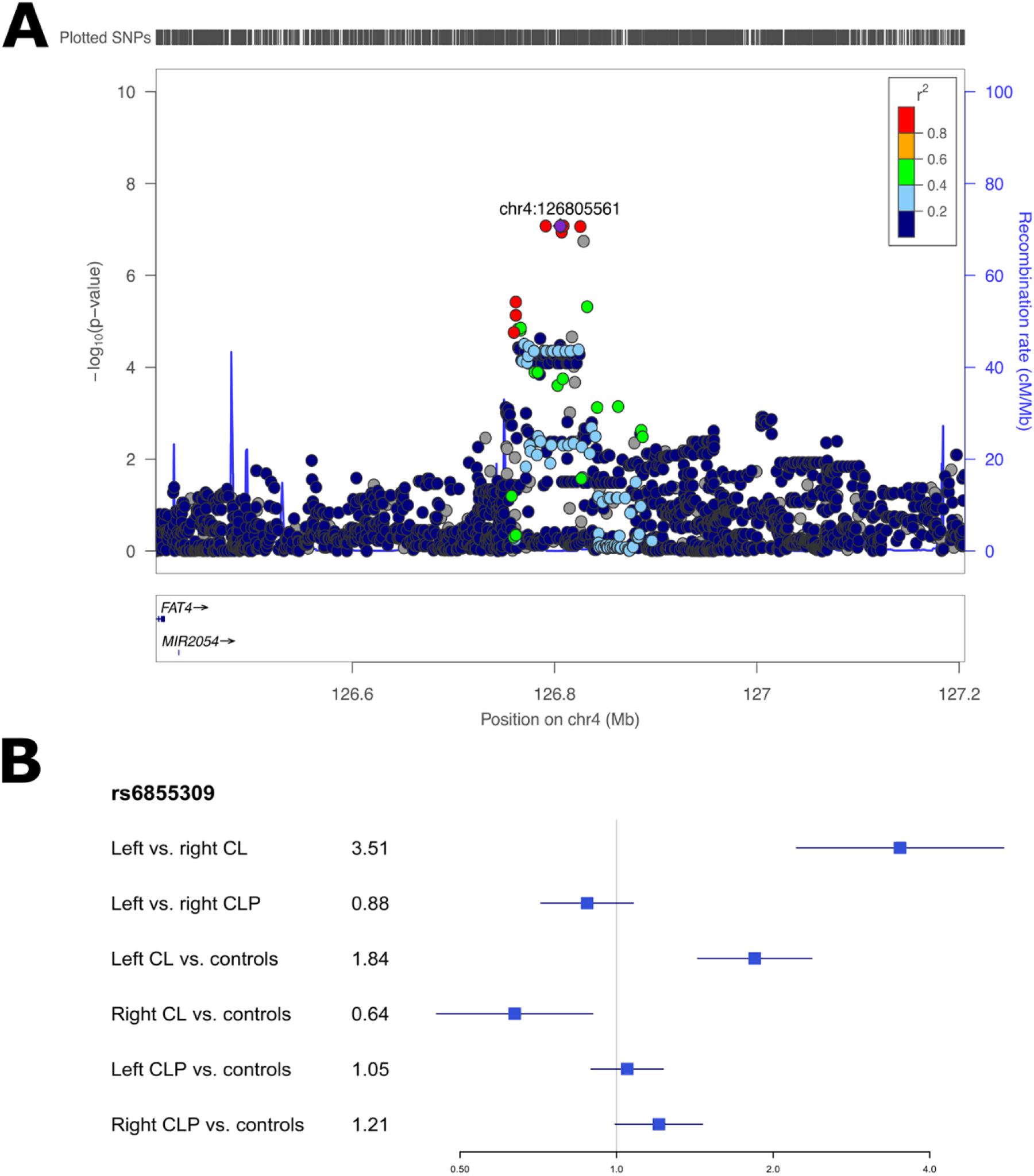
Regional association plots showing –log_10_(p-values) for the suggestive peak at 4q28 in the modifier analysis in cleft lip (A). Plots were generated using LocusZoom (R. J. Pruim et al., 2010).The recombination overlay (blue line, right y-axis) indicates the boundaries of the LD block. Points are color coded according to pairwise LD (r^2^) with the index SNP. The odds ratio for this locus in each of the modifier and subtype specific loci were also compared (B).

**Figure 5:**
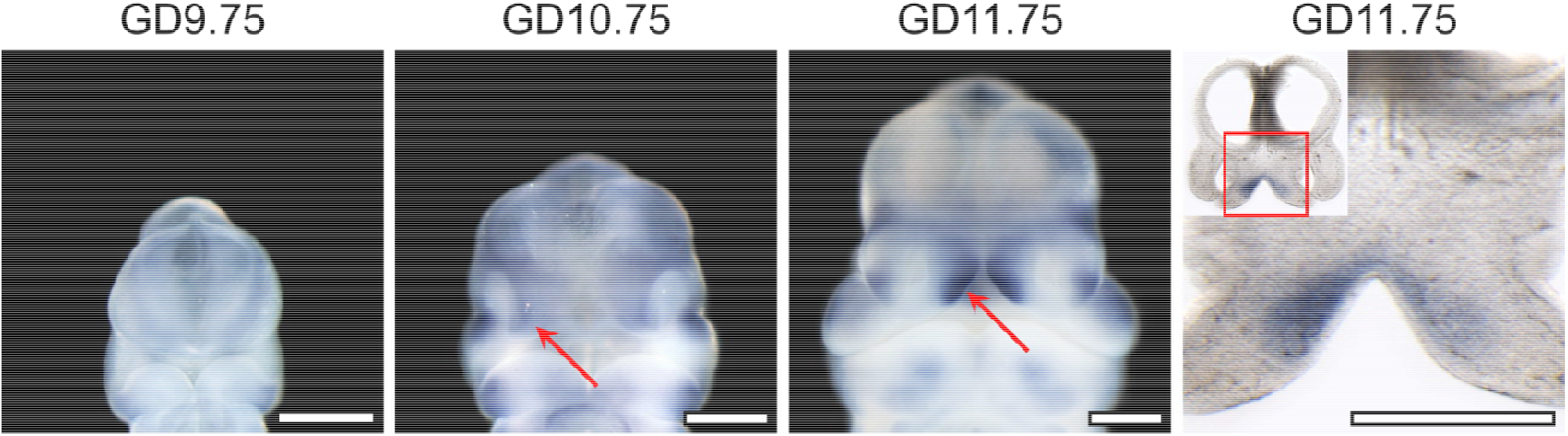
*Fat4* Expression in the Medial Nasal Processes During Orofacial Morphogenesis. Staining for *Fat*4 by *in situ* hybridization in mouse embryos at indicated gestational days (GD) demonstrates its expression in the medial nasal process (red arrow) that form the median aspect of the upper lip. A coronal section at GD11.75 and magnified view of the boxed region shows that *Fat4* is specifically expressed in the mesenchyme of the medial aspects of the medial nasal processes. Scale bar = 500 μm.

### Genetic interaction with *FAT4*

We were also interested in whether this potential modifier locus interacted with regions near known OFC risk loci, so we tested for interaction between the lead SNP at the *FAT4* gene and SNPs within 50kb of previously associated risk loci. In LCL, two separate regions in 1p31.1 showed significant evidence of interaction with rs6855309. The first locus had a protective interaction (Lead SNP: rs1405051; OR = 0.53; p = 2.48 × 10^−5^), which was also seen in LCLP (OR = 0.66; p = 3.87 × 10^−5^), but not in RCL or RCLP (p > 0.05; Table S10). The second locus, 1 MB away, appeared to increase risk for LCL only (Lead SNP: rs12034480; OR = 2.43; p = 3.89 × 10^−9^), with no significant interaction in any other subtype. This suggests, while the exact mechanism by which modifiers act is not known, modifier loci can interact with known OFC loci to increase or decrease risk for specific subtypes of OFCs.

### Epigenomic context of results and gene set enrichment test

Given the modifier and subtype-specific loci were largely distinct, we tested whether these loci were enriched for distinct functional regions involved in human facial development. Because GWAS-associated SNPs are primarily noncoding, we annotated each SNP based on functional segments based on epigenetic and histone modifications in human embryonic craniofacial tissues (Wilderman et al., 2018). The results were similar across timepoints throughout craniofacial development (4.5-8 weeks post conception), so here we report on Carnegie Stage 15 (CS15) as a representative analysis (Figure S6; Table S11). The top SNPs (p < 1 × 10^−3^) had different enrichment and depletion in several functional segments (Figure 6). For example, subtype-specific loci in LCL and RCL were enriched in regions of strong transcription (LCL: OR = 1.97, p = 2.24 × 10^−39^; RCL: OR = 1.24, p = 0.001), but subtype-specific loci in LCLP and RCLP were depleted in this region (LCLP: OR = 0.66, p = 1.81 × 10^−7^; RCLP: OR = 0.45, p = 6.43 × 10^−17^). Additionally, LCL was enriched in repressed polycomb regions (OR = 2.09, p = 4.97 × 10^−6^) whereas SNPs were depleted in this region in LCLP (OR = 0.23, p = 0.0002). For these potential modifier loci, there were also differences across cleft type, with modifier loci in the LCL-RCL analysis being depleted in enhancer regions (OR = 0.63, p = 0.0002), while modifier loci in LCLP-RCLP analysis were enriched in this region (OR = 1.23, p = 0.02). This further supports the hypothesis that the genetic underpinnings of cleft subtypes are distinct and may involve different biological mechanisms. Finally, we tested whether any of these SNPs in the modifier analyses or any of the subtype specific analyses were enriched in genes previously associated with other laterality disorders, however, no gene set showed statistical significance (Table S12).

**Figure 6:**
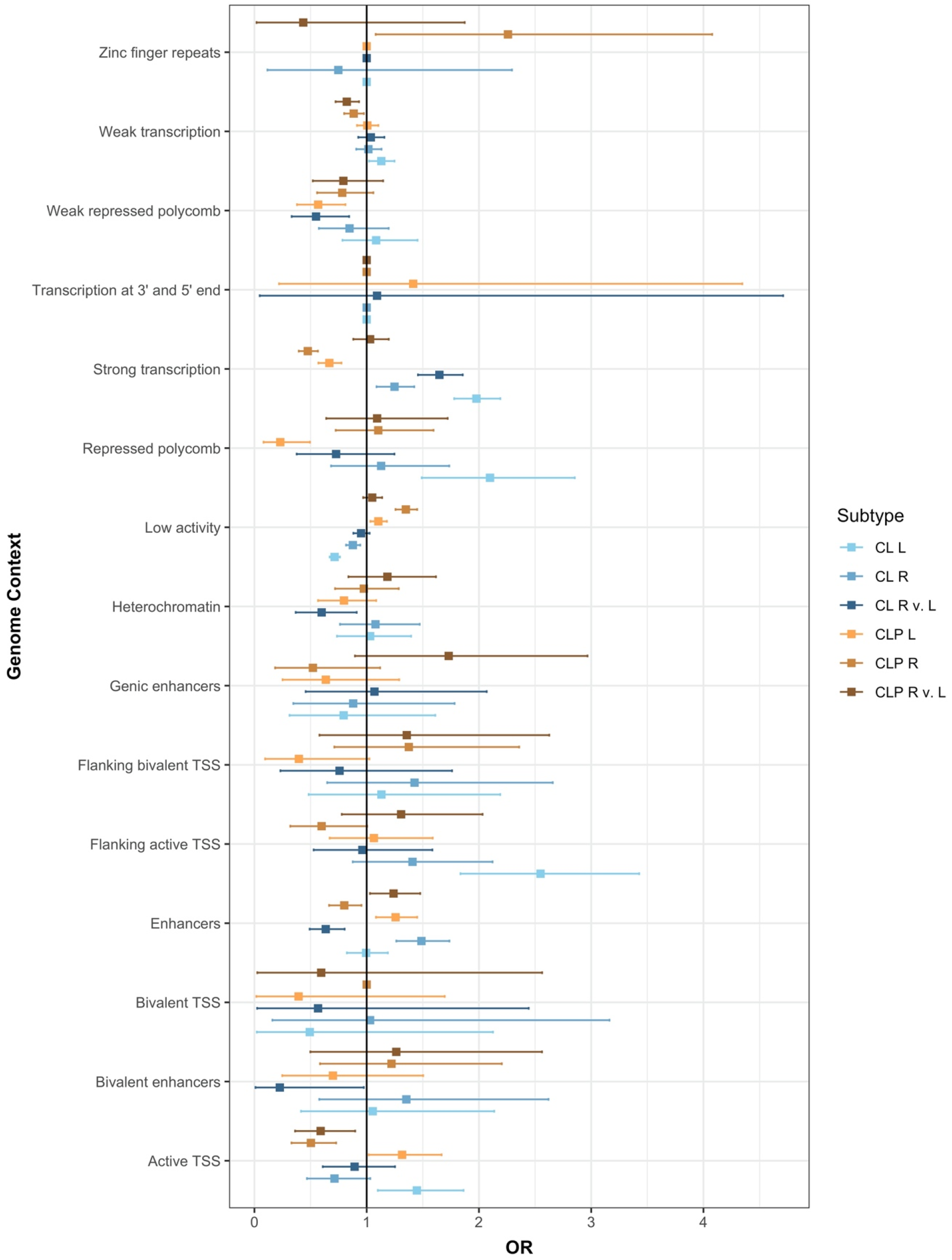
Enrichment of the top SNPs associated in either the CL modifier analysis, CLP modifier analysis, and each subtype analysis (p < 1 × 10^−3^) were tested in each functional region defined during craniofacial development (CS15).

### Discussion

OFCs are a heterogeneous group of craniofacial birth defects with multiple distinct subtypes, however, while many studies have been done to better understand the genetic underpinnings of OFCs overall, much less work has been done to understand the genetics leading to the individual subtypes, such as left versus right unilateral defects. To better understand potential underlying genetic factors controlling risk to distinct unilateral OFC subtypes, we conducted a modifier-based approach to LCL, RCL, LCLP, and RCLP within a multiethnic case-control study. While no locus reached genome-wide significance, we identified a suggestive locus at 4q28, which increased the odds of a left unilateral cleft lip over right unilateral cleft lip. We were unable to replicate this locus in the GENEVA sample although this may be due to technical or statistical challenges stemming from different SNP arrays and imputation panels, sample size, and/or population differences. For example, the replication sample was predominantly of Asian ancestry (73.6%), and the associated SNPs at the 4q28 locus had lower allele frequencies in Asian populations (MAF: 2-15%) than either European populations (MAF: 42-60%) or Latin American populations (MAF: 24-33%) that made up most of the discovery sample. Therefore, issues with statistical power could explain why this locus did not replicate.

Nonetheless, several lines of evidence support the biological plausibility of this locus being involved in craniofacial development and laterality. The closest protein coding gene, *FAT4*, is located approximately 400kb from the associated region. *FAT4* encodes a protocadherin involved in planar cell polarity, a process involved in tissue organization and left-right differentiation (Saburi et al., 2008). Mutations in *FAT4* have been linked to both autosomal recessive Hennekam (Alders et al., 2014; Ivanovski et al., 2018) and Van Maldergem syndromes (Cappello et al., 2013; Ivanovski et al., 2018). Hennekam syndrome is a disorder resulting from malformations in the lymphatic system and can cause facial dysmorphism (Lister Hill National Center for Biomedical Communications, 2020), while Van Maldergem syndrome is characterized by intellectual disabilities, craniofacial abnormalities, and other skeletal defects. *FAT4* was previously linked to OFCs in a previous study reporting a burden of missense variants in *FAT4* associated with OFCs in a large, extended Syrian pedigree (Holzinger et al., 2017). Additionally, we found that *Fat4* is expressed in mouse embryos in the mesenchyme of the medial nasal processes at gestational day 10.75 and gestational day 11.75. At this developmental stage, the paired medial nasal processes are a precursor to the upper lip and undergo concerted outgrowth and fusion with the adjacent lateral nasal and maxillary processes, closing the upper lip. Previous work has found that disrupting the proliferation of mesenchymal cells in the medial nasal processes results in cleft lip (Everson et al., 2017). Therefore, *FAT4* is expressed at the proper time and place to be involved in the differentiation between left-sided and right-sided cleft lip. Further analysis of the expression and function of *FAT4* in this region could help elucidate the gene’s possible involvement in the development of left versus right cleft lip.

This is one of the first genome-wide studies to analyze genetic differences associated with the side of unilateral clefts; it supports a growing body of evidence that genetic differences underly the phenotypic heterogeneity of OFCs (Carlson, Standley, et al., 2017; Carlson, Taub, et al., 2017; K. U. Ludwig et al., 2016). In addition to finding candidate genetic modifiers for right unilateral and left unilateral CL and CLP, comparisons of the effect sizes suggest that the effects are not shared between CL and CLP. This suggests that the genetic modifiers for left vs. right-sided clefts may be distinct in CL and CLP. Supporting this, we found no new modifiers in the analysis of CL and CLP combined (CL/P), despite the larger sample size which should have increased statistical power. Furthermore, the modifier loci in these analyses showed opposite enrichments in some functional regions, as defined by the Epigenomic Atlas of Human Craniofacial Development (Wilderman et al., 2018), with the modifiers in CLP being enriched in enhancer regions, while modifiers in CL were depleted in enhancer regions. This suggests different loci may be associated with RCL vs. LCL compared to RCLP vs. LCLP, but that there may also be heterogeneity in the mechanisms behind how these modifiers act.

We also conducted a subtype-specific GWAS comparing the four case groups with controls. These analyses would have less statistical power to detect loci differing between any two subtypes, but should find loci associated with either overall OFC risk or risk of one particular cleft subtype. The loci that reached genome-wide significance in these analyses were loci that were already associated with OFCs in previous analyses (Carlson et al., 2019; Huang et al., 2019; Leslie, Carlson, et al., 2016; Yu et al., 2017). However, when we compared the odds ratios for all the suggestive SNPs in the subtype-specific analyses and tested if the confidence intervals of these odds ratios overlapped for genetic variants in different subtypes, some of the suggestive loci showed different effects between OFC subtypes. As with modifier analyses, the top loci in each subtype-specific analysis were enriched in different functional regions. For example, the loci associated in left unilateral CL and right unilateral CL were enriched in regions of strong transcription, whereas these regions were depleted in the top SNPs from the analyses of left unilateral CLP and right unilateral CLP. This suggests the potential for some heterogeneity in the biological mechanisms behind distinct OFC subtypes, which warrants further study.

This study is not without limitations, which include small sample sizes for some subtypes. Previous studies have identified population-specific association signals, but the small sample sizes precluded further stratified, population-specific analyses. Because of the diversity of these samples, we had to account for population structure by adjusting for a large number of principal components of ancestry, which could further reduce statistical power. Given that the 4q28 locus did not replicate in a cohort with more participants of Asian ancestry, there is possibility population-specific genetic modifiers have yet to be discovered. Larger samples from each ancestry group could provide more statistical power and the ability to identify population-specific modifiers.

In conclusion, we utilized a modifier-based approach to develop a better understanding of the genetic differences of LCL, RCL, LCLP, and RCLP. This approach allowed us to identify a candidate region on 4q28 that increases the odds for a LCL over RCL. *FAT4*, the gene closest to this region, has previously been reported in an extended pedigree with nonsyndromic OFCs, and the location of this gene’s expression during murine development suggests *FAT4* may have some role in craniofacial development. We initially set out to determine if there was a genetic basis to the preponderance of left-sided unilateral clefts shared by CL and CLP. By analyzing CL and CLP separately, we found many of the genetic associations’ corresponding functional enrichments were distinct between CL and CLP and between the sides of these clefts. These results highlight the highly heterogenous nature of OFCs and how potential genetic mechanisms for OFC pathogenesis may be overlooked by combining all OFC subtypes for analysis based on epidemiological similarities.

## Data Availability

Original data used in this study is available at dbGaP (phs000774.v2.p1, phs000094.v1.p1, https://www.ncbi.nlm.nih.gov/gap/)

## Acknowledgments

The authors thank the dedicated field staff, collaborators, and participating families for their important contributions to this study.

